## Supplementary figures and images for "Enrichment of *SARM1* alleles encoding variants with constitutively hyperactive NADase in patients with ALS and other motor nerve disorders"

### Figure 2 - figure supplement 1

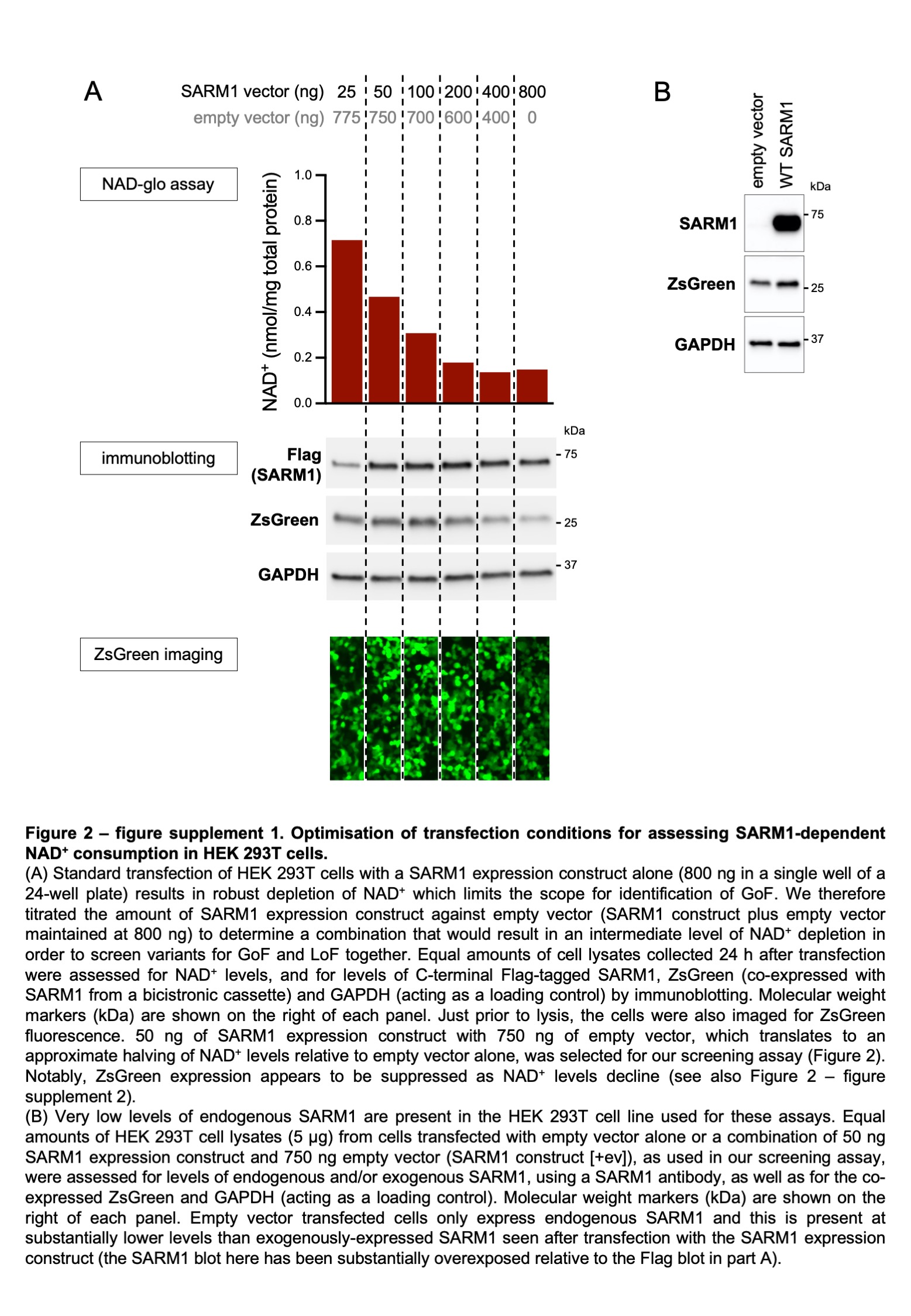

### Figure 2 - figure supplement 2

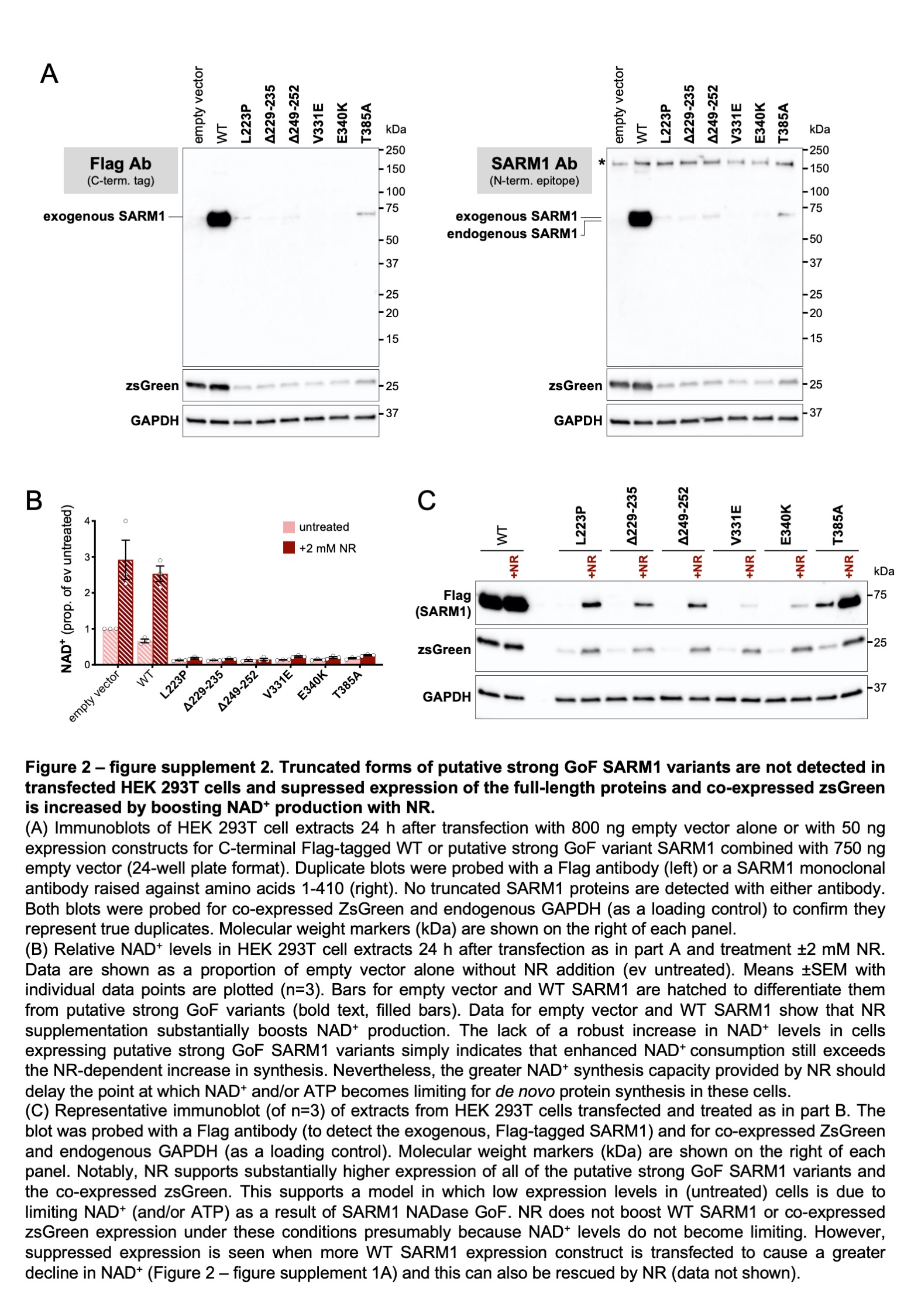

### Figure 2 - figure supplement 3

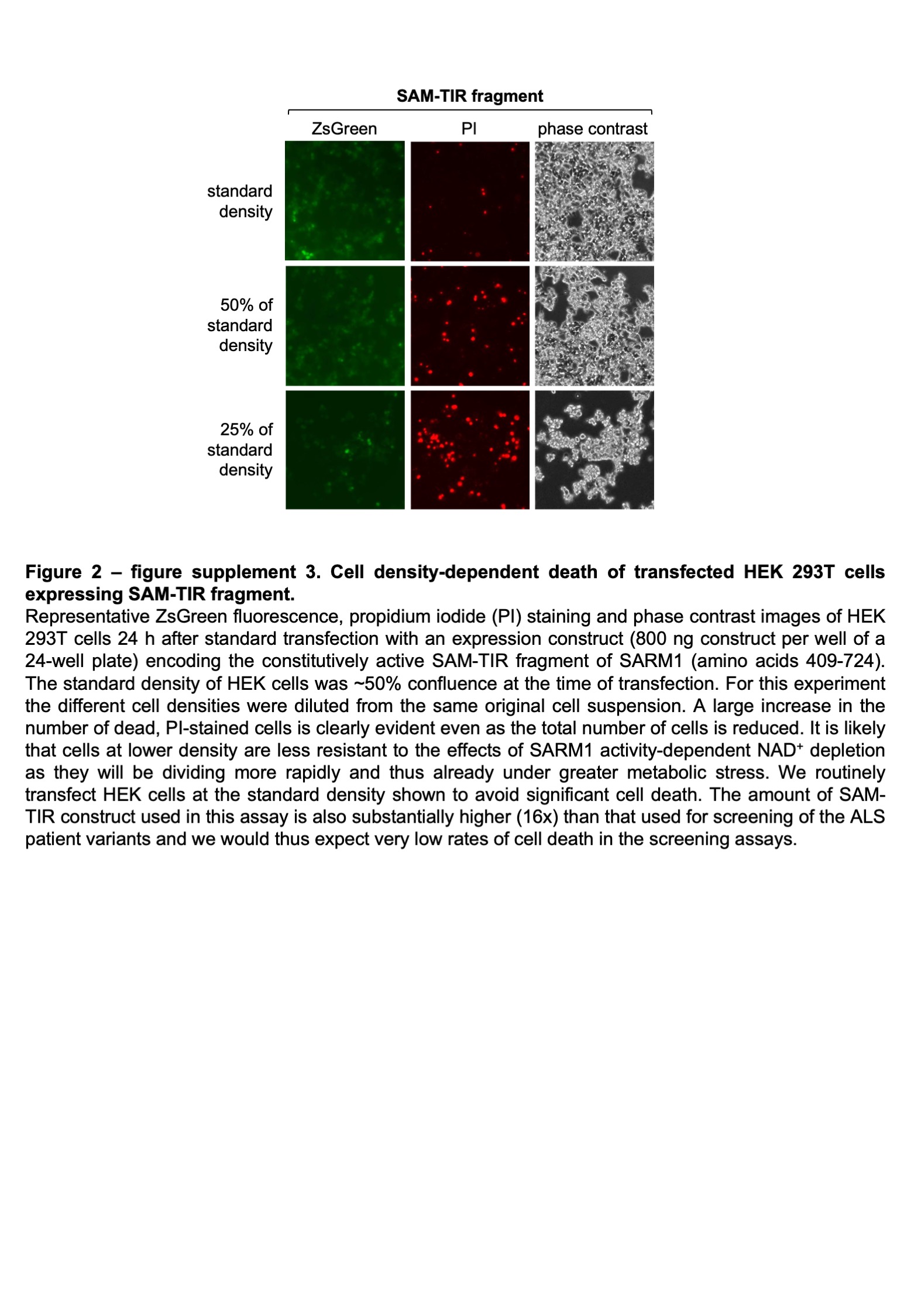

### Figure 3 - figure supplement 1

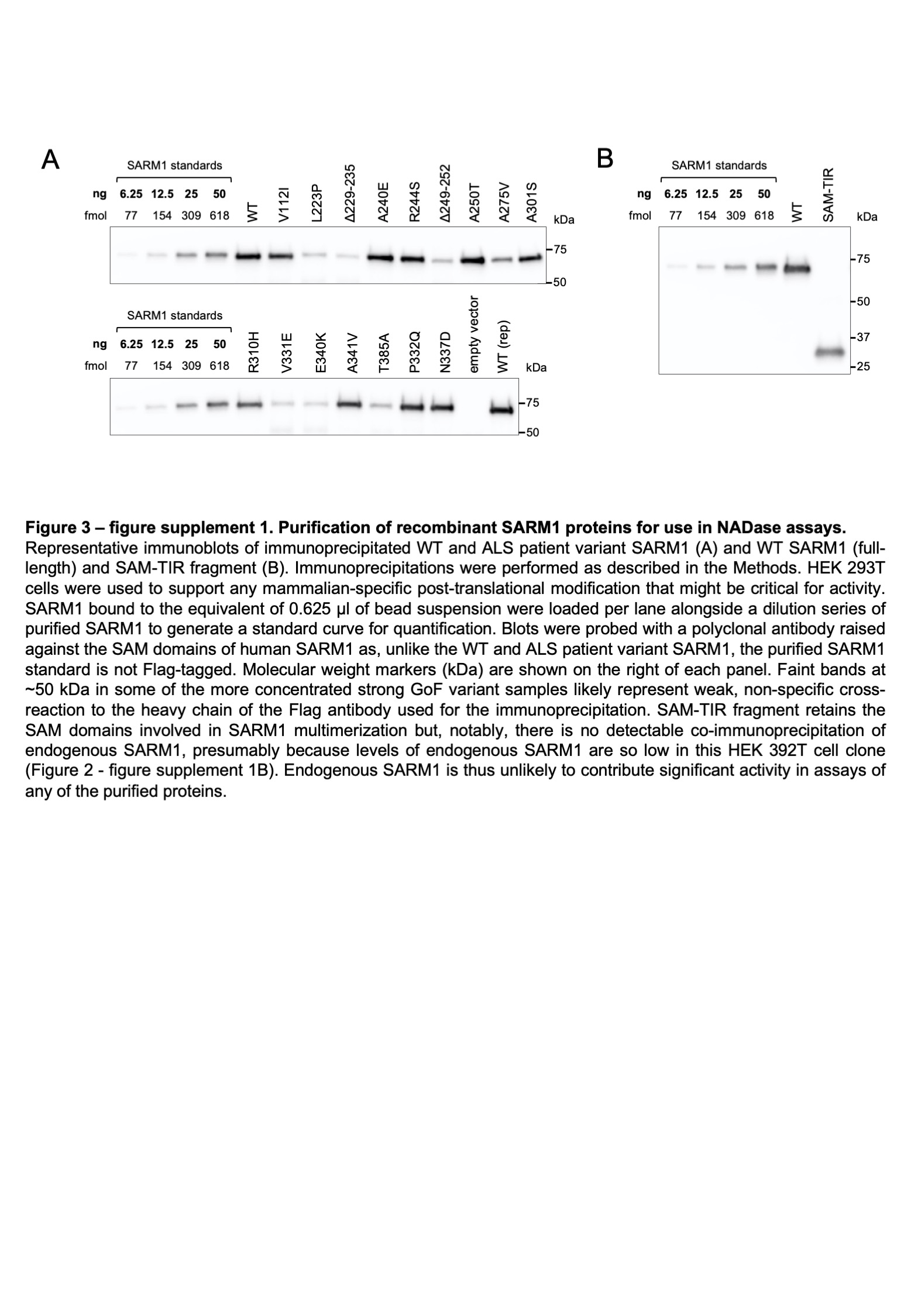

### Figure 3 - figure supplement 2

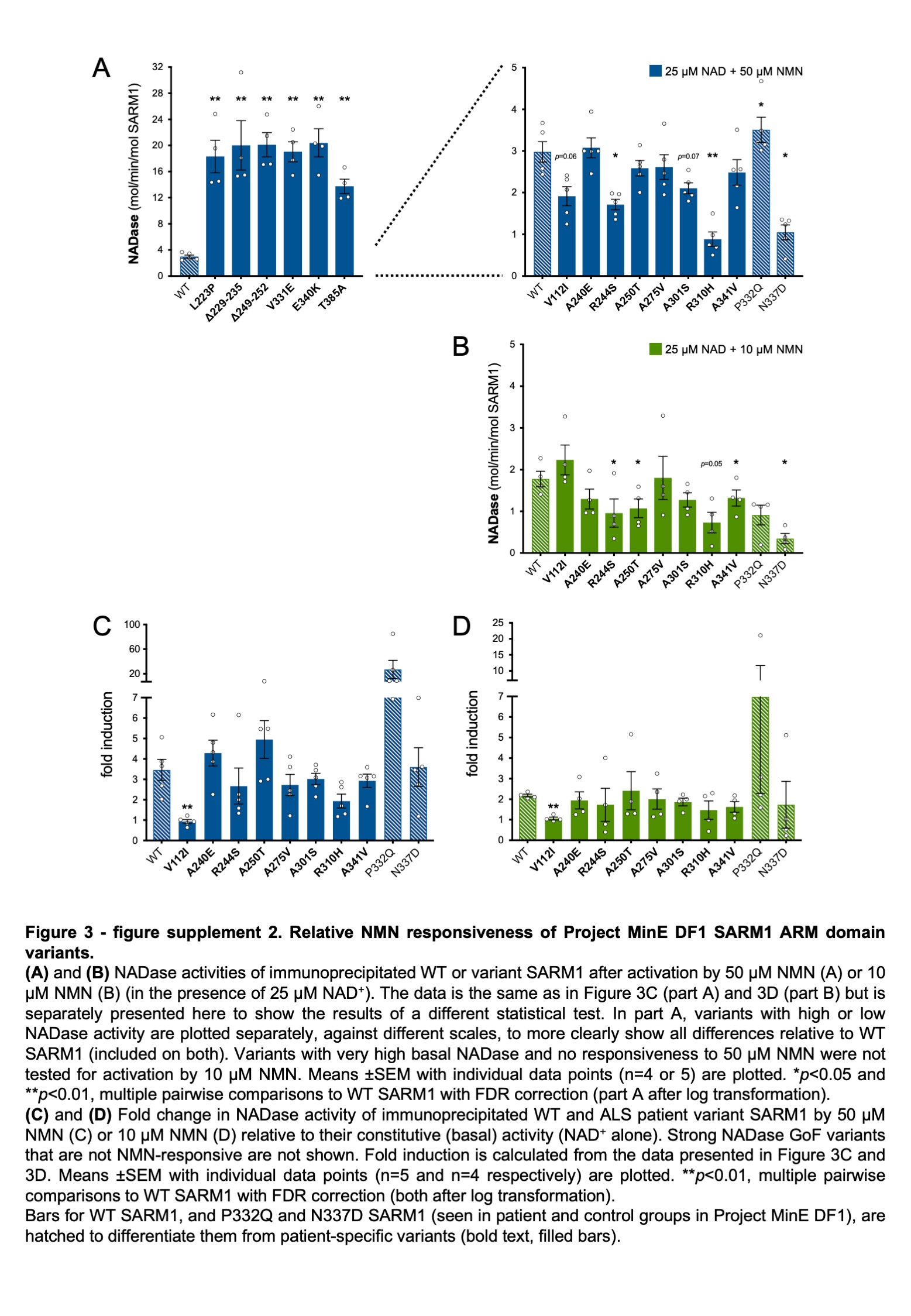

### Figure 6 - figure supplement 1

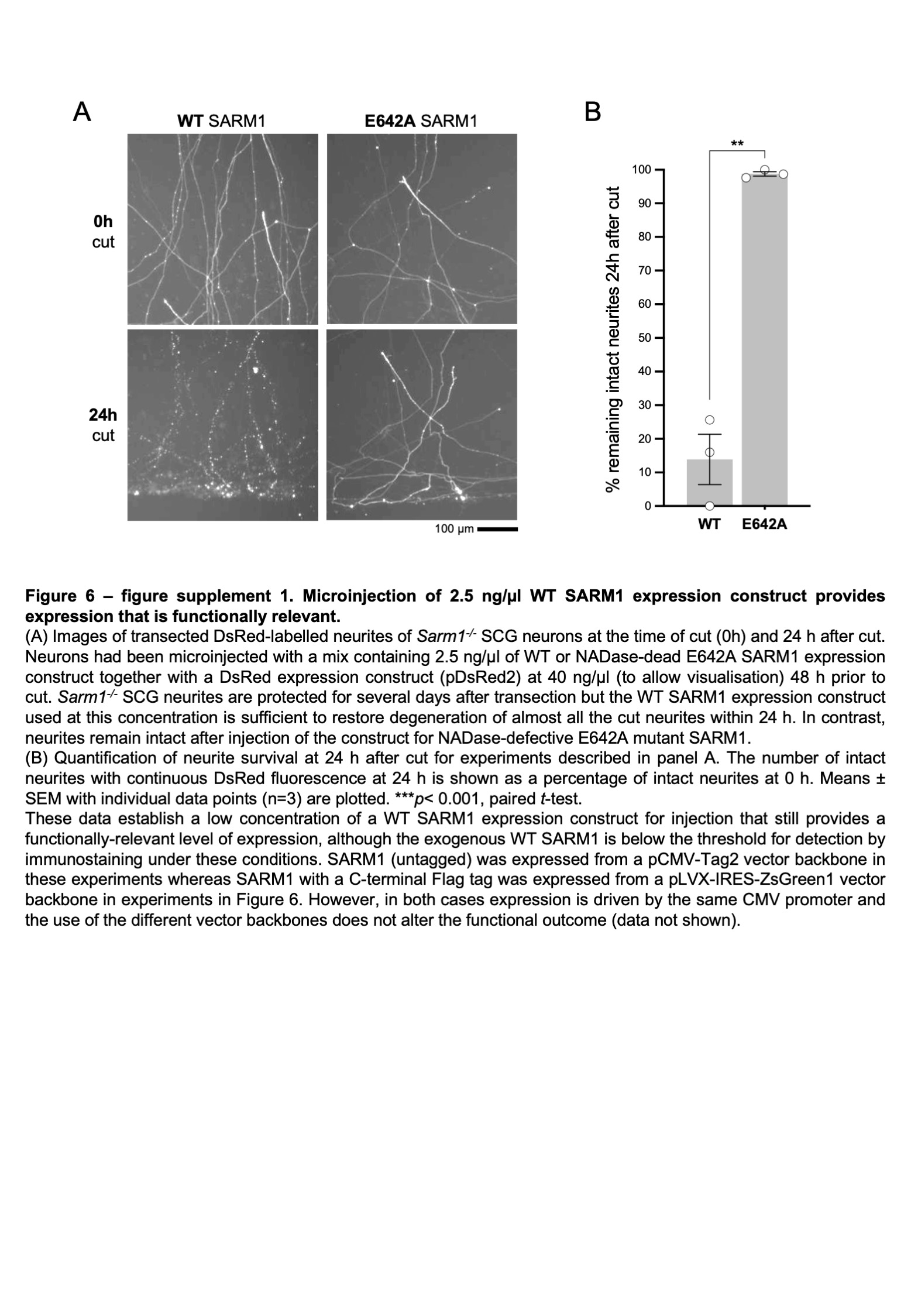

### Figure 6 - figure supplement 2

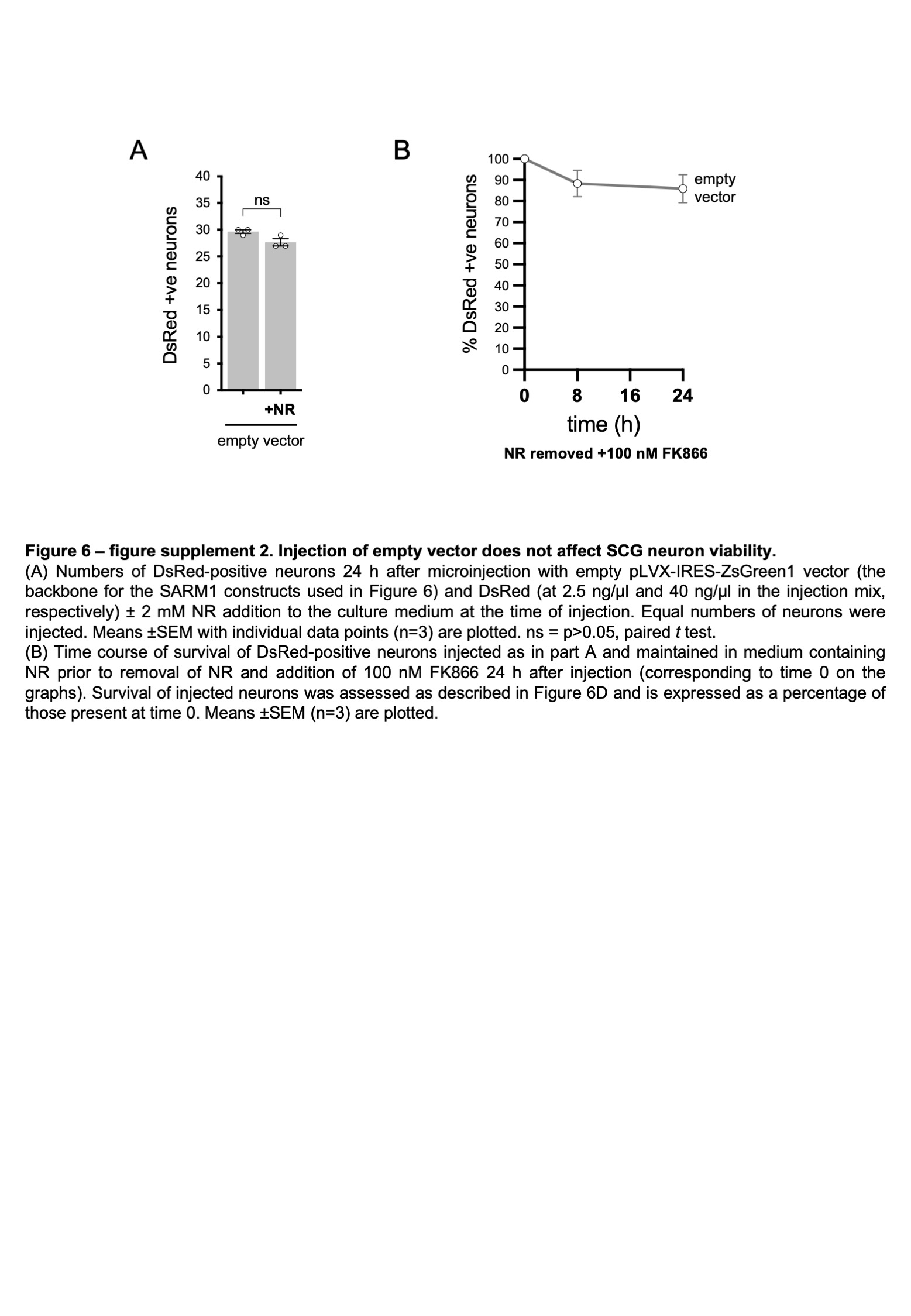
